# Changing COVID-19 vaccine eligibility could reshape disease burden for all

**DOI:** 10.64898/2026.04.27.26351870

**Authors:** Soren L. Larsen, Pamela P. Martinez, Ayesha S. Mahmud

**Affiliations:** Department of Demography, University of California Berkeley, Berkeley, CA, USA; Department of Microbiology, University of Illinois Urbana-Champaign, Urbana, IL, USA; Department of Statistics, University of Illinois Urbana-Champaign, Urbana, IL, USA; Carl R. Woese Institute for Genomic Biology, University of Illinois Urbana-Champaign, Urbana, IL, USA

**Author notes:** To whom correspondence should be addressed, RM310 Social Science Bldg., Berkeley, CA 94720, United States.

## Abstract

COVID-19 vaccine recommendations have changed in the United States over the past year, raising questions about the likely impact on disease dynamics. While older adults are most at risk of severe COVID-19 outcomes and therefore experience the greatest direct benefits of vaccination, limiting vaccination to only this age group could worsen outcomes in this higher-risk population through indirect effects. Here, we leveraged survey data on age-specific vaccine uptake, and surveillance data from Illinois to calibrate a transmission model accounting for contact and vaccination rates across age. We simulated four years of COVID-19 transmission under three scenarios: vaccination uptake across all ages, vaccination of only those aged 5+, and vaccination of only those aged 65+. Our model suggests that removing those under 65+ from eligibility could increase hospitalizations by 23-27% across age groups, signaling that vaccination coverage among younger adults and children substantially mitigates COVID-19 hospitalization burden for older populations. Moreover, while children would experience the greatest per capita increase in infections, older adults would experience the greatest per capita increase in hospitalizations. The full scope of consequences for disease burden may not be observed until after the first year of the policy change. These findings highlight the long-term consequences of deprioritizing young adults and children from vaccination campaigns, and show that gaps in vaccine-induced immunity can shape outcomes in other age groups. This underscores the importance of considering indirect vaccine impacts when developing policy.

## Introduction

In 2025, the United States underwent a high-profile federal policy shift in age-based recommendations for COVID-19 vaccination [1]. This included revoking approval of Pfizer’s Comirnaty vaccine for children under 5 and updating under-65 eligibility for COVID-19 immunizations to include only those with at least one underlying risk factor for severe COVID-19 outcomes [2, 3]. At the same time, several states developed their own policy positions, such as New Mexico’s expanded eligibility guidelines [4], or moved to create new governing bodies, such as the newly formed West Coast Health Alliance [5]. Recommendations from physician groups further complicated this policy picture. For example, the American Academy of Pediatrics recommended that all children 6-23 months receive the 2025-26 COVID-19 vaccine as part of the initial series or an additional updated dose [6], inconsistent with federal guidelines. This heterogeneous policy landscape raises questions about the potential population-level impacts of age-related vaccine eligibility on disease dynamics over time, and requires a consideration of both direct and potential indirect vaccination impacts.

The safety and efficacy of COVID-19 immunizations have been clearly established [7]. Given the stark age-gradient in the risk of severe outcomes from COVID-19 [8], older adults experience the greatest direct benefits of vaccination. However, the potential indirect impacts of vaccinating younger adults and children should not be discounted, and under some conditions prioritizing younger people for vaccination may be a mortality-minimizing strategy [9]. Being surrounded by vaccinated contacts can provide indirect protection, depending on the infection and transmission-blocking properties of the vaccine; this would be consistent with herd immunity, where “the return on every individual immunized is more than one individual” [10]. In turn, clustering of unvaccinated contacts is likely to compound transmission risks. This may be facilitated through known age-based homophily [11], or through younger adults living with their children, especially in light of high household secondary attack rates documented in previous work [12]. Previous studies have evaluated eligibility scenarios for updated vaccine formulations in a single season, for example the US Scenario Modeling Hub in 2024-25 [13] and 2025-26 [14], finding substantial indirect impacts for adults over 65 when vaccinating the entire population with updated formulations. These studies emphasize the importance of vaccinating younger individuals, and provide crucial insights into short-term impacts of updated vaccine campaigns. However, the pre-existing vaccination landscape is likely to buffer vaccine policy impacts in the short-term; while dimensions of protection conferred by vaccines wane over time and decline in response to variant replacement, protection against severe disease has been shown to remain high even 6 months after vaccination [15].

Sustained changes in vaccine policy - resulting in a declining proportion of vaccinated individuals over the long-term due to births, deaths, and waning of vaccine-induced immunity - may drive greater increases in severe disease burden over time than what has been estimated for the short-term. Alternatively, the impact of lower vaccination rates may become attenuated over subsequent years, as the proportion with natural immunity increases and a new endemic equilibrium is established. The importance of characterizing this nonlinear system is underscored by previous work exploring re-emergence of vaccine-eliminated infectious diseases in the United States, finding that long-term case burden and time to endemicity were highly variable by pathogen and degree of declining vaccination coverage [16]. In light of these open questions, there is a need for studies that explore potential long-term COVID-19 dynamics both in the presence of child and young-adult vaccination, and in its absence.

To address this gap, we implemented an age-stratified transmission model to explore differences in disease dynamics under a range of age-based vaccination scenarios: (1) vaccination of all ages, (2) vaccination of those age 5+, and (3) vaccination of those age 65+. To capture existing seasonal transmission dynamics and account for heterogeneous vaccination, contacts, and severity across age groups, we calibrated this model to data from Illinois during the 2024-25 respiratory disease season, supplementing with survey data as a benchmark for vaccination and contact patterns. We then simulated four years of COVID-19 transmission under each vaccination scenario, finding that while restricting vaccination to ages 5+ would have limited impacts, restricting to ages 65+ could substantially increase disease burden for all groups. Importantly, while per capita increases in infections were highest among children, the greatest hospitalization impacts were seen in adults 65+, and this was consistent across a range of scenarios with differing levels of vaccine efficacy against infection and transmission. Disease dynamics were least impacted during the first year of a policy change, then stabilized at consistently higher weekly hospitalizations in years two through four, suggesting that policy impacts may be underestimated by shorter term studies. These results have potential implications for the long-term consequences of declining vaccine-induced protection among younger people, which may substantially increase disease burden for all, and especially for those over 65.

## Methods

### Survey sample

We previously fielded a survey of Illinois residents during the 2024-25 respiratory disease season, asking questions about health and health-seeking behaviors related to respiratory pathogens [17]. Using data for 8, 963 survey respondents (Table S1), we estimated the age-stratified prevalence of ever having received a COVID-19 primary series vaccination and multiple additional doses. Details of the survey data and analysis can be found in the supplementary materials.

### Transmission model

To explore vaccine impacts on COVID-19 disease burden in the context of heterogeneous vaccination behavior, we used an age-stratified Susceptible-Exposed-Infectious-Hospitalized-Recovered-Susceptible model that accounts for contact structure within and between age groups, as well as vaccination patterns by age (Figure S1). We simulate the outbreak in a population of 12,812,508 individuals, where 5.1% are children under 5, 16.4% are children 5-18, 60.9% are adults between 19-64, and the remaining 17.5% are 65+, reflecting the Illinois population size and structure [18].

Informed by CDC surveillance data from 2024 [19], in all simulations, we assume that 5% of the population starts as recently recovered (*R*_*age*_ and *RV*_*age*_) and 0.1% of the population is distributed across infected (*I*_*age*_, *IV*_*age*_) and hospitalized (*H*_*age*_, *HV*_*age*_) compartments (Tables S2). The remainder of the population begins in the susceptible compartments (*S*_*age*_ and *SV*_*age*_), moving to the exposed compartments (*E*_*age*_, *EV*_*age*_) at a rate ω_*age*_(*t*), which is the time-dependent force of infection stratified by age and vaccination status. Vaccination reduces the risk of infection by *V E*_*i*_, the vaccine efficacy against infection. Once exposed, individuals move at a rate ε to either the infected (*I, IV*) or hospitalized (*H, HV*) compartments; the hospitalization rate *η*_*age*_ is dependent on their age and reduced, if vaccinated, by vaccine efficacy against severe disease *V E*_*s*_. Infected individuals will eventually recover at a rate *γ*, while hospitalized individuals may recover or die according to an age-specific infection fatality ratio *σ*_*age*_. Once recovered, immunity from infection wanes at a rate *ω*. Vaccine-induced immunity wanes at a rate *ω*_*v*_, moving individuals from *RV*_*age*_ to *R*_*age*_, and *SV*_*age*_ to *S*_*age*_. In addition to infection- and vaccination-related movement between compartments, individuals in each age group age at rates *α*_*age*_, and those leaving 65+ compartments are replaced by new births in the *S*_0−4_ compartment. Model equations for unvaccinated and vaccinated adults, aged 19 to 64, are provided below as an example:

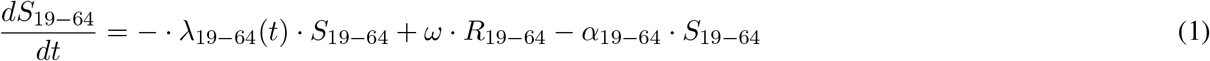

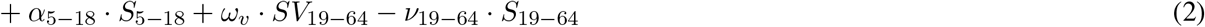

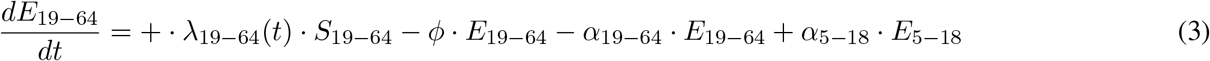

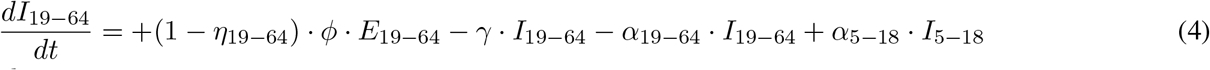

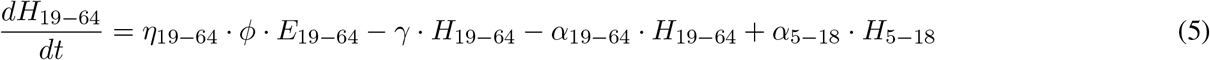

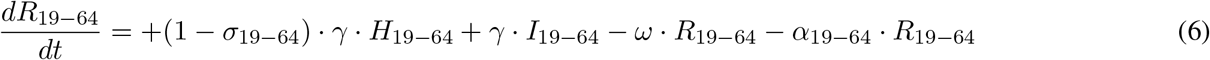

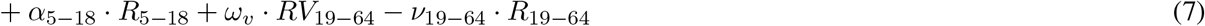

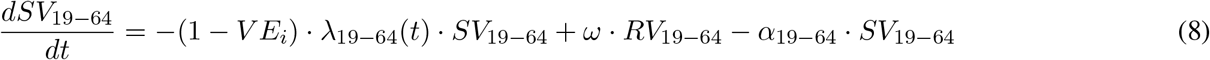

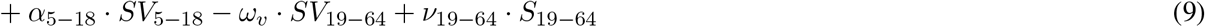

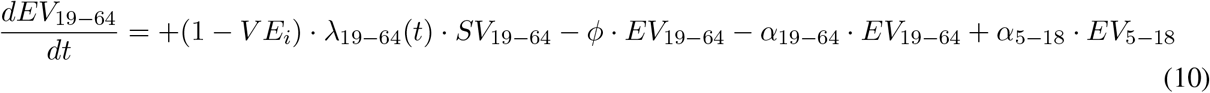

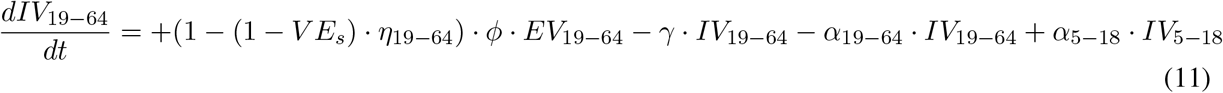

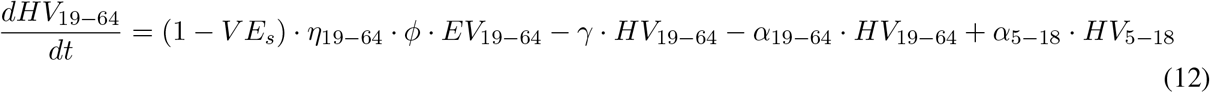

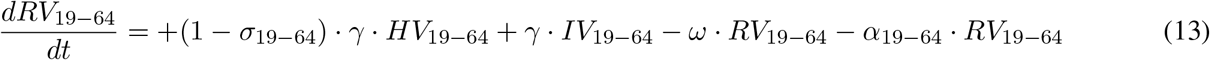

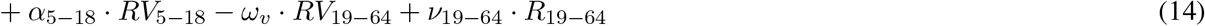

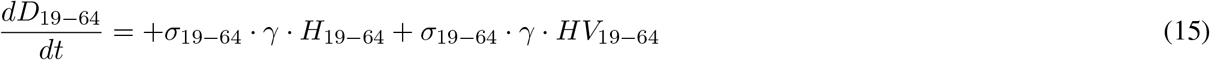

We parameterized this model with estimates of the risk of severe disease using data provided by the Illinois Department of Public Health [20] and age-stratified infection fatality ratios weighted to Illinois population structure [8, 21]. Parameter values used in simulations are given in Table S3. We used the *DeSolve* package [22] in R to find numerical solutions.

### Vaccination

When initializing the model, individuals were distributed across vaccinated and unvaccinated SEIRS compartments based on survey proportions (Tables S4, S5), assuming that 25% of those reporting prior vaccination would have current vaccine-induced immunity at the start of simulations.

To capture ongoing vaccination throughout simulations, we first fit age-stratified type-III functional response curves to monthly vaccination data in Illinois - reported by the CDC during the 2024-25 respiratory disease season [23] - following the methods detailed in [24]. Functional response parameters are provided in Table S6. Then, these functions were used to estimate daily vaccination rates for each age group and day of the year, where vaccination moves individuals from *S*_*age*_ to *SV*_*age*_, and from *R*_*age*_ to *RV*_*age*_.

We implemented three possibilities about the potential indirect impacts of vaccination: (1) vaccination reduces transmission risk only (main text), (2) vaccination reduces infection risk only, and (3) vaccination reduces both infection and transmission risks. We assumed an overall vaccine efficacy of 45% against severe disease, informed by estimates for older adults during the 2024-25 season [25]. This overall efficacy is not conditional on infection. Therefore, in scenarios where vaccination does not block infection, we adopted *V E*_*s*_ = 45%. In scenarios where vaccination does block infection, we assumed a conservative vaccine efficacy of 34% against infection, which is lower than 2023-24 estimates of efficacy against *symptomatic* infection [26] - and set *V E*_*s*_ = 17% so that the combined efficacy against severe disease, accounting for *V E*_*i*_, was 45% overall. In transmission-blocking scenarios, vaccine efficacy against transmission for infectious individuals was set at 22%, informed by [27]. Reductions in transmission are also supported by the finding that vaccinated individuals have a shorter duration of infectious virus shedding than those who are unvaccinated [28].

### Force of infection

The time-dependent transmission rate *β* (*t*) is generated from a cyclic cubic spline basis, using the function *cSplineDes()* from the package *mgcv* [29]. The basis takes six magnitude parameters *µ*_*i*_, each corresponding to a two-month period, to capture seasonality in transmission throughout the year. While transmission could also change over time as a result of introduction of new variants, exploring variants of concern is beyond the scope of this analysis. The time-dependent force of infection for each age group, ω_*age*_(*t*), is calculated from the transmission rate *β* (*t*) and the rate of contact with infectious members of the population, as follows:

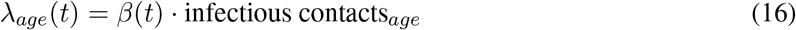

### Contacts

We calculated age-stratified contact rates with the *socialmixr* package [30], using UK POLYMOD rates [31] reweighted to the Illinois population age distribution [18]. The infectious contacts for an individual in a given age group are as follows: where *τ* is the reduction in an infectious individual’s transmission potential conferred by vaccination.

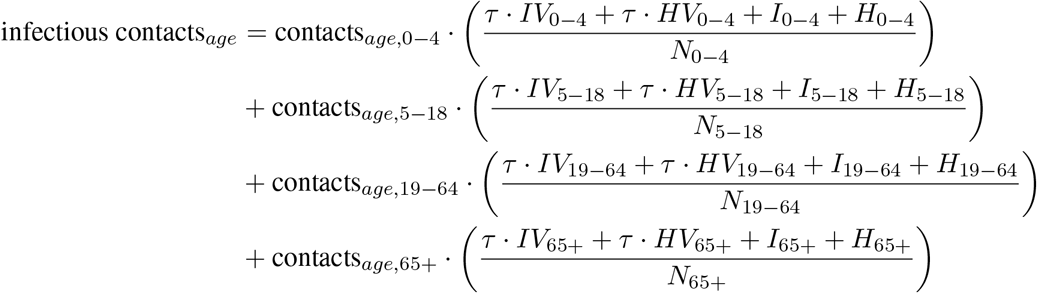

### Calibration

Using the method of maximum likelihood with the *bbmle* package [32], we fit the model to weekly, population-level COVID-19 deaths in Illinois, reported by CDC, from March 2024 to March 2025 [33] (Figure S2). We used mortality data for calibration due to its accessibility and reliability; mandatory reporting of hospitalizations ended in May 2024 [34], and estimates of hospitalization fatality rates are based on data from earlier years [8, 20]. For fitting, the modeled and observed data were normalized by the maximum deaths observed during this period. For each set of hypotheses about indirect vaccination impacts, we obtained maximum likelihood estimates for the six magnitude parameters that define seasonal transmission. We calculated log-likelihood using the normal probability distribution *dnorm()* in base R. After calibrating models to data, we profiled the magnitude parameters defining transmission by rerunning the maximum likelihood estimation function at a sequence of focal parameter values, allowing all other parameters to vary. Confidence intervals (95%) are computed using a spline function to estimate the parameter values *p* for which *ll*_*p*_ = *ll*_*ref*_ − 1.92, where *ll*_*ref*_ is the maximum log-likelihood [35]. Maximum likelihood estimates and corresponding confidence intervals are provided in Tables S8-S10.

### Scenarios

We explored three scenarios of vaccination coverage in the population: (1) a reference scenario, where age-stratified vaccination patterns continue as observed in the 2024-25 season (“Vaccinate all-ages”); (2) a scenario where only those age 5+ are vaccinated (“Vaccinate 5+”), and (3) a scenario where only age 65+ are vaccinated (“Vaccinate 65+”). We also simulated an intermediate scenario where vaccination is primarily restricted to those age 5+ but some parents continue vaccinating their children regardless of official recommendation, which is included in the supplement. Age-specific vaccination coverage parameters for each scenario are provided in Table S7.

The mechanisms driving COVID-19 seasonality are complex and an area of ongoing investigation [36]. While we calibrated seasonal forcing parameters to capture two annual waves, we also conducted a sensitivity analysis to explore a wider range of seasonal dynamics. To explore uncertainty in model outcomes across the parameter space, we first sampled 4,000 sets of seasonal transmission parameters from a Latin hypercube of the 95% confidence intervals. Then, we filtered for those parameter combinations whose average value matched the average value of the maximum likelihood estimates to three decimals. This ensured that overall transmission was similar across replicates while allowing for variation in wave timing and magnitude throughout the year. This resulted in 94 replicates for the transmission-blocking scenario, 101 for the infection-blocking scenario, and 101 for the transmission- and infection-blocking scenario. When estimating cumulative infections, hospitalizations, and deaths in each scenario, we report the value at maximum likelihood parameter estimates, as well as the 2.5 and 97.5 percentiles of sensitivity analysis trajectories. To estimate the difference in infections, hospitalizations, and deaths compared to reference scenarios, we paired trajectories by their transmission parameter values and reported both the maximum likelihood estimate and 2.5 and 97.5 percentiles of paired differences. We emphasize analysis of hospitalizations over infections or deaths; while mortality rates are overall low, hospitalizations leading to those deaths remain a greater burden while providing information on severe outcomes that infection incidence does not.

For each scenario and set of transmission parameters, we simulated one year of transmission under all-ages vaccination (the reference scenario), starting at the initial conditions in Table S2. After one year, we updated the vaccination coverage functions and simulated four years of transmission under the new policy. This four-year period is the focus of our analysis. In the reference scenario where all ages are eligible for vaccination, daily vaccination rates for the 2024-25 season are repeated in subsequent years. In counterfactual scenarios where some age groups are ineligible for vaccination, we adjust the potential maximum vaccinated *V*_*m*_ to 0 throughout years 1-4 of the simulations, and recalculate daily vaccination rates accordingly.

## Results

To understand the potential impacts of changing vaccination policy, we calibrated an age- and vaccination-stratified transmission model (Figures 1A, S1-S2) to a year of disease dynamics under an all-ages vaccination policy (“Year 0”), then simulated alternative vaccination scenarios for an additional four years (“Years 1-4”; Figures 1B-C). If an all-ages vaccination policy is maintained in Years 1-4, our model estimates 1.90 hospitalizations per 100, 000 in children under 5 (2.5-97.5 percentile [1.14, 1.99] in sensitivity analysis), 17.59 [10.71, 18.46] in older children (5 − 18), 307.29 [183.75, 323.01] in younger adults (19 − 65), and 407.07 [239.86, 429.86] among older adults (65+) (Figures 1C, S3-S5). In analyses that follow, we take this as the reference scenario. Our model suggests that restricting vaccination to those aged 5+ in Years 1-4 would have a negligible impact on population-level COVID-19 dynamics (Figures 1B, 2). However, restricting vaccination to those 65+ could substantially increase hospitalization burden, with the greatest impacts observed in Years 2-4 (Figure 2) - despite a buildup of more recovered individuals in the population in the same time period (Figure S6).

**Figure 1:**
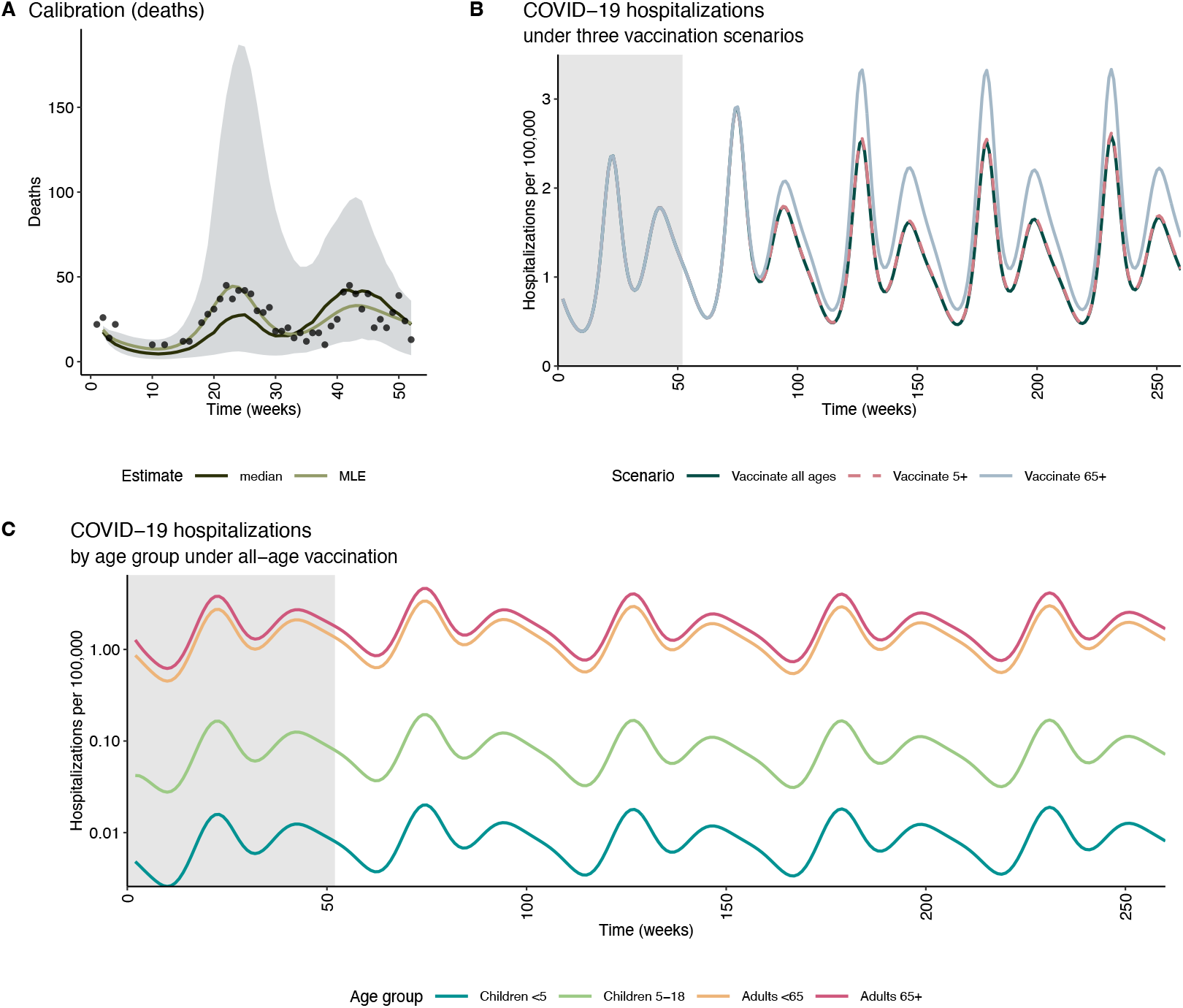
(A) Simulated (lines) and observed (points, [33]) weekly deaths in Year 0 under all-ages vaccination, assuming that vaccines reduce transmission potential for infectious individuals. Lines represent the model outcomes at maximum-likelihood transmission parameter estimates from model calibration, as well as the median from transmission parameter sensitivity analysis (94 trajectories). Ribbons represent the weekly 2.5 and 97.5 percentiles across trajectories. (B) Model-simulated weekly hospitalizations per 100, 000 in Year 0 (all-ages vaccination policy; grey shading) and Years 1-4 under alternative scenarios (“Vaccinate all-ages”, “Vaccinate 5+”, and “Vaccinate 65+”). We assume that vaccination reduces the transmission potential of infectious individuals, and simulations represent maximum likelihood parameter estimates. (C) Age-stratified weekly hospitalizations per 100, 000 in Years 0-4, under all-ages vaccination. Initial conditions and model parameters can be found in Tables S2-S3, and distribution of vaccination coverage in Tables S4 and S5.

**Figure 2:**
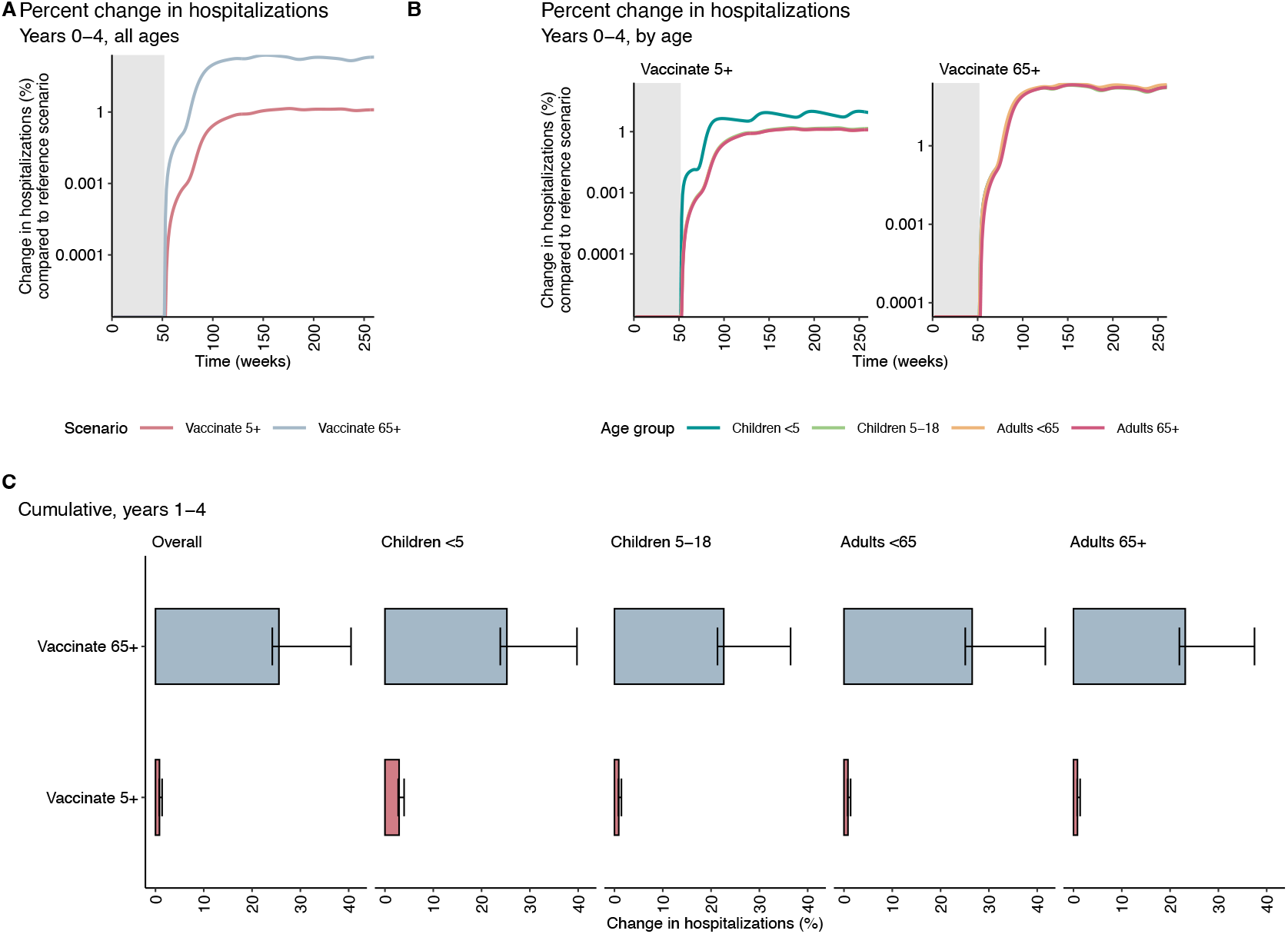
(A) Percentage increase in model-simulated weekly hospitalizations per 100,000 over years 0-4 in alternative scenarios (“Vaccinate 5+” and “Vaccinate 65+”) compared to the reference scenario where all ages continue to receive vaccinations. We assume that vaccination reduces the transmission potential of infectious individuals, and simulations represent maximum likelihood parameter estimates. (B) Age-stratified percentage increases in model-simulated weekly hospitalizations per 100,000 over years 0-4 in alternative scenarios (“Vaccinate 5+” and “Vaccinate 65+”) compared to the reference scenario where all ages continue to receive vaccinations. (C) Percentage increase in model-simulated hospitalizations attributable to COVID-19 over Years 1-4, at maximum likelihood parameter estimates. Two alternative eligibility scenarios are compared to the reference scenario with all-ages vaccination. Bars represent 2.5 and 97.5 percentiles of 94 paired trajectories from sensitivity analysis, with parameters sampled from their 95% confidence intervals. Initial conditions and model parameters can be found in Tables S2-S3, and distribution of vaccination coverage in Tables S4 and S5.

In particular, if only those age 5+ were eligible for vaccination in Years 1-4, our model suggests a small percent increase in COVID-19 hospitalizations across groups relative to all-ages vaccination, ranging from 0.82% [0.78%, 1.38%] (adults 65+) to 2.92% [2.74%, 3.97%] (under five years; Figure 2C). Framed in per capita terms, these increases translate to 0.06 [0.04, 0.07] additional hospitalizations per 100, 000 among children *<* 5, up to 3.36 [3.27, 3.53] per 100, 000 among adults 65+. In other words, despite observing the greatest per capita increases in infections among children 5 − 18 and adults *<* 65 (Figure S7), we find that older adults would experience the greatest per capita increase in COVID-19 hospitalizations (Figure S8) and deaths (Figure S9) if vaccination is age-restricted. In the scenario where only those 65+ are eligible for vaccination, we find greater percent increases in hospitalizations relative to all-ages vaccination, ranging from 22.63% [21.33%, 36.46%] (children 5-18 years old) to 26.54% [25.11%, 41.68%] (adults *<* 65 years old). The greatest per capita change in hospitalizations is again seen for older adults, with an additional 94.19 [89.85, 99.50] hospitalizations per 100, 000 relative to an all-ages vaccination policy (Figure S8), translating to a 23.14% [21.92%, 37.48%] increase (Figure 2C). These findings signal considerable indirect impacts of vaccinating younger people for COVID-19 despite the substantially lower risk of severe outcomes in this population.

Finally, while we assumed that vaccination reduces the risk of transmitting as an infected individual, there is uncertainty in the extent of this mechanism, and it is also possible that vaccines could mitigate the risk of becoming infected as a susceptible individual. To address this uncertainty, we first implemented the assumption that vaccination blocks both infection and transmission, and recalibrated the model accordingly. Under this assumption, our model estimates older adults to have 369.27 [268.20, 405.55] COVID-19 hospitalizations per 100, 000 under all-ages vaccination, with an increase of 1.97% [1.81%, 2.74%] if only ages 5+ are vaccine-eligible, and 55.50% [51.12%, 76.50%] if only ages 65+ are vaccine-eligible (Figure S10). Next, we tested the assumption that vaccination only blocks infection, without blocking transmission (Figure S11); our model estimates older adults to have 389.75 [283.62, 428.22] hospitalizations per 100, 000 under all-ages vaccination, with an increase of 1.30% [1.19%, 1.81%] if only ages 5+ are vaccine-eligible, and 36.36% [33.46%, 50.09%] if only older adults are vaccine-eligible. These simulations highlight the potential for considerable population-level impacts on hospitalizations among older adults even if vaccine efficacy against transmission is limited, and especially if there is vaccine efficacy against infection.

## Discussion

Indirect impacts of vaccination can be hard to intuit from measures of vaccine efficacy alone, but mathematical models that capture nonlinear disease dynamics are well-suited to explore these impacts. Here, we used survey data on vaccination and surveillance data on disease prevalence for the state of Illinois to inform a transmission model that accounts for heterogeneous vaccination patterns across age groups. Using this model, we simulated five years of COVID-19 transmission under an all-ages vaccination policy and compared with alternative scenarios where, after the first year, vaccination is restricted to those age 5+ or those age 65+. In these scenarios, we found consistently higher disease-related hospitalizations among all age groups, and especially among older adults despite unchanged coverage in this age group. Despite observing the greatest per capita increases in infections among younger adults and children in counterfactual scenarios, adults 65+ consistently had the greatest change in per capita hospitalizations and deaths among any age group. The bulk of impacts on disease burden were observed after the first year under the new policy, underscoring that the consequences of policy changes may not be readily observable in the short-term. Our results complement prior work showing indirect benefits from universal vaccination with updated boosters [13, 14], while adding to the existing literature by exploring longer-term dynamics.

Our findings suggest that all-age COVID-19 vaccination mitigates a substantial burden of disease, especially for adults 65+, and that the long-term consequences of changing vaccine policy may not be evident early on. This also suggests that policies driving a long-term increase in un- or under-vaccinated younger people could result in markedly increased hospitalizations among older adults. Serious adverse events from COVID-19 vaccination are rare [37], making it exceedingly unlikely that indirect increases in COVID-19 hospitalizations or deaths would be outweighed by any decrease in vaccine-related adverse events. Importantly, data from the National Immunization Survey suggests 17.5% of adults and 8.9% of children received an updated COVID-19 vaccine as of February 21 in the 2025-26 season [38] compared to 22.7% and 11.9% by the same time in the season prior [39]; policy alone does not explain low rates of updated booster uptake, and interventions to address vaccine hesitancy remain important.

We stress that our simulations are not intended to quantitatively predict future disease burden, but rather to qualitatively explore indirect impacts of vaccination coverage patterns across age groups, and several limitations should be considered when evaluating our findings. First, our model was benchmarked with data from a single state; further work is needed to ensure that these trends are generalizable to the broader U.S. population. We relied on self-reported vaccination status to initialize the model, which may have inaccuracies due to recall bias or social desirability bias. While we accounted for age, we did not explicitly incorporate household structure or homophily by vaccination status, which may result in underestimating transmission risk. Whereas absolute estimates of hospitalization burden should be treated with caution, we are confident in the validity of cross-scenario comparisons, which yielded qualitatively consistent findings across a range of assumptions and transmission parameters. Second, our modeling analysis represents a set of simplified scenarios intended to isolate the effect of age-stratified vaccination coverage on disease dynamics. Therefore, vaccine efficacy and timing are held constant across seasons, and viral evolution is not considered. While beyond the scope of this study, recent work suggests that these factors contribute to the biannual SARS-CoV-2 waves observed throughout the United States [36], and the timing of both vaccination [14, 24] and waning immunity [40] have been shown to be important contributors to disease dynamics. Despite these limitations, our model can provide qualitative insights into possible future dynamics. Key directions for future work include examining geographic variation in the possible impact of changing vaccination policies, exploring more granular age-stratification in direct and indirect vaccine impacts, and incorporating heterogeneity in population risks across sociodemographic groups.

In conclusion, despite relatively modest risks of severe disease for younger populations, existing COVID-19 vaccination patterns are likely averting a substantial burden of disease among adults older than 65 years. As vaccine coverage declines among younger individuals in the US in coming years, we encourage further work in this area to inform public policy decisions and healthcare preparedness.

## Acknowledgements

The authors would like to thank the Illinois Department of Public Health for generously providing SARS-CoV-2 surveillance data.

## Funding statement

The survey presented here was funded by a grant from SHIELD Illinois. S.L.L. and A.S.M. received funding for this research from the National Institute of General Medical Sciences of the National Institutes of Health under award number R35GM156856.

## Author Contributions

All authors conceptualized the publication. S.L.L. drafted the manuscript; A.S.M. and P.P.M. reviewed and edited the manuscript.

## Competing Interests

The authors report no competing interests.

## Data availability

Data and code used to generate results in the manuscript will be made available in a permanent Zenodo repository upon publication.

## Declaration of generative AI and AI-assisted technologies in the manuscript preparation process

During the preparation of this work the authors used Claude AI to generate a compartmental model diagram from model equations. After using this tool, the authors reviewed and edited the content as needed and take full responsibility for the content of the published article.

## Notes

### Competing Interest Statement

The authors have declared no competing interest.

### Author Declarations

IRB of the University of Illinois Urbana-Champaign IRB waived ethical approval for this work (reference number IRB42-1168)

### Summary of Updates

This version of the manuscript has been updated to expand this work from a brief report to a full article. We have substantially updated our model structure and assumptions, and added new analyses. The central conclusion, that restricting vaccination to older adults could increase disease burden for all ages and especially for those 65+, is unchanged.

