## Supplementary material for "Changing COVID-19 vaccine eligibility could reshape disease burden for all": SI

#### Supplementary Text

##### Illinois Survey

During the 2024-25 respiratory disease season, we conducted a statewide survey in Illinois on health and health-seeking behaviors related to respiratory pathogens [17]. The survey was implemented in two waves (11/24 - 12/24; 02/25- 03/25) using the Qualtrics platform and respondents were recruited through Dynata, an online panel provider (Dynata, Shelton, CT, USA, https://www.dynata.com). Our survey study protocol received an IRB exemption determination from the University of Illinois Urbana-Champaign IRB (reference number IRB42-1168). All participants were provided information on the study procedure, risks, and benefits, as well as points of contact. By taking part in the study, participants confirmed that they had read the information in the consent form, had the opportunity to ask questions, and voluntarily agreed to take part in the study. We screened respondents for those over 18 and living in Illinois, and excluded those who did not meet this criteria. For both waves, we used a reCaptcha threshold of $0.5$ to remove responses that were identified as likely to be bots. In Wave 2 only, we required successful completion of a Captcha question at the beginning of the survey. Across waves, we removed responses with duplicate *psid* values, which were unique to respondents.

Our final sample (Table S1) consisted of survey respondents who provided responses to the following questions:

1. Which of the following best describes your yearly combined household income from all sources?
2. What is the highest educational level you have completed?
3. Which of the following best describes your race? (Please select all that apply.)
4. Are you of Hispanic or Latino origin (excluding origins in Spain)?
5. What is your year of birth?
6. Have you ever been vaccinated against COVID-19?
7. If you have been vaccinated against COVID-19, did you receive: (Only primary series; Primary series and one booster; Primary series and multiple boosters)

We combined questions 4 and 5 into a single variable for race and ethnicity. We use “Hispanic or Latino” in this work to refer to individuals of any single racial identity who report Hispanic or Latino ethnicity. Terms such as “White” refer to those individuals who report one racial identity and do not report Hispanic or Latino ethnicity, unless otherwise stated. Additionally, if respondents reported at least one child in the home, we required that they responded to:

1. Has your child ever been vaccinated against COVID-19?
2. If your child has been vaccinated against COVID-19, did your child receive: (Only primary series; Primary series and one booster; Primary series and multiple boosters)

After these selection steps, 8,963 respondents remained. We used raked weighting to adjust the survey sample to improve sample representation of Illinois population distributions by race and ethnicity, age, income, and education. The weights were generated using Census data from Illinois [18] and the *weights* [41] and *anesrake* [42] packages in R with a maximum weight size of 5. Weighted and unweighted sample characteristics can be found in Table S1. For the purposes of this analysis, vaccinated individuals were those that reported receiving a primary series vaccine and multiple additional doses. We calculated weighted vaccination rates for children under $5$, children $5-18$, adults $19-64$, and adults $65+$.

#### Supplementary Figures

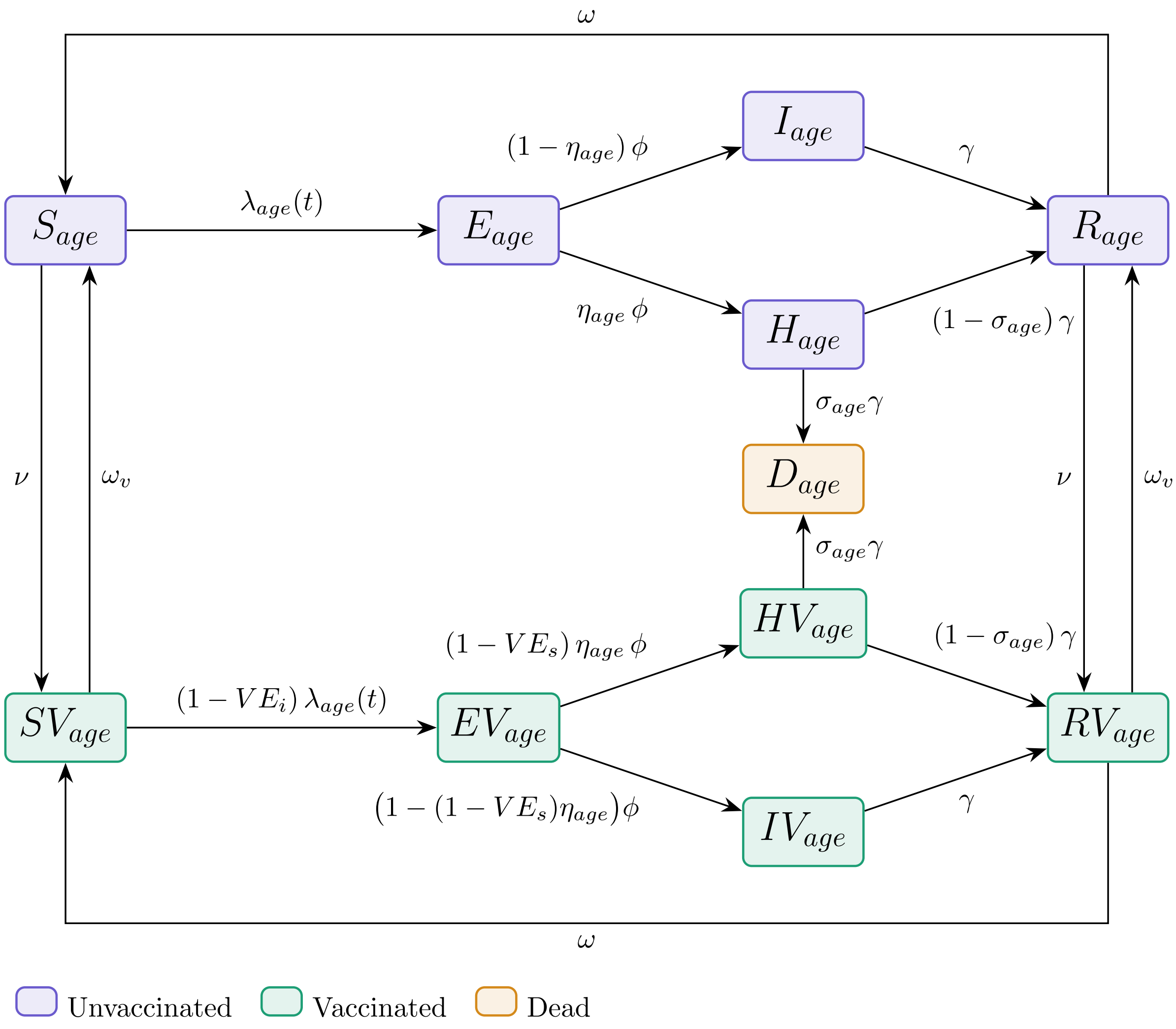


Figure S1: Compartmental model diagram for a given age group. Flows due to aging are excluded for clarity.

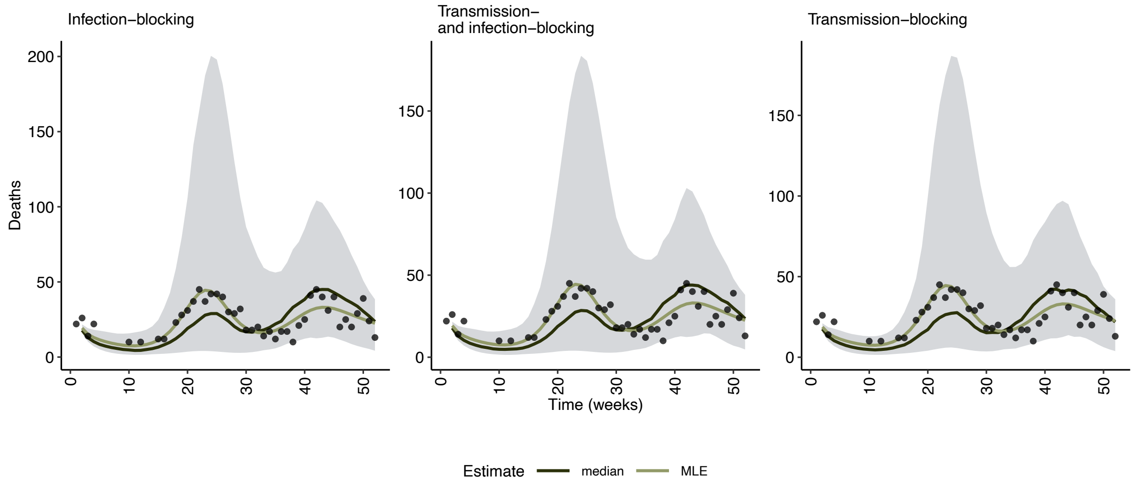


Figure S2: Simulated (lines) and observed (points) weekly deaths in Year 0 under all-ages vaccination, assuming that vaccines block infection risk for susceptible individuals, transmission potential for infectious individuals, or both. Lines represent the model outcomes at maximum-likelihood transmission parameter estimates from model calibration, as well as the median from transmission parameter sensitivity analysis (101, 94, and 101 trajectories for infection-blocking, transmission-blocking, and infection- and transmission-blocking scenarios respectively). Ribbons represent the weekly 2.5 and 97.5 percentiles across trajectories.

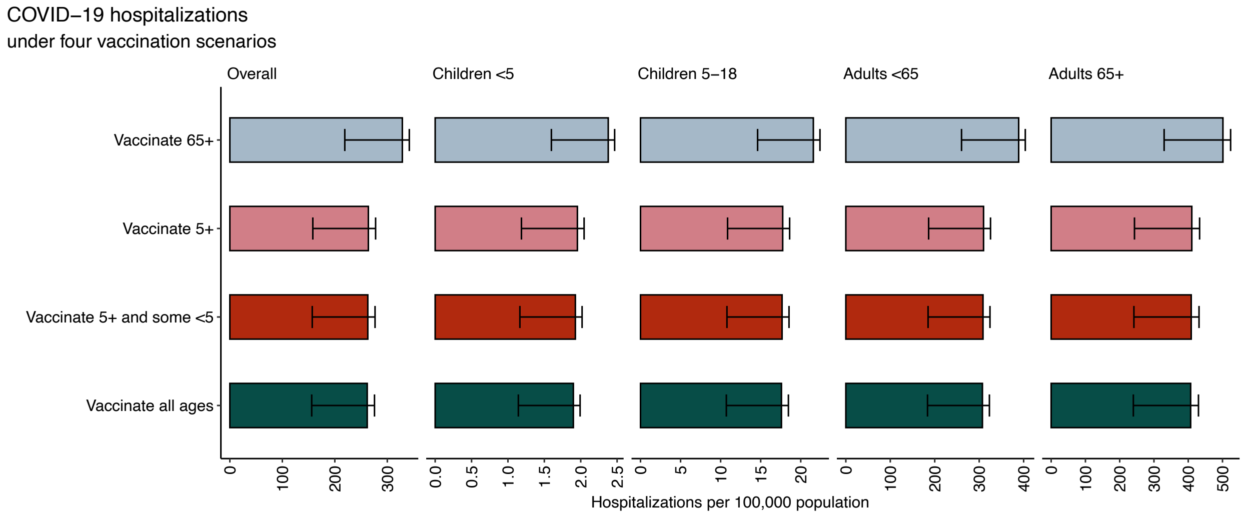


Figure S3: Model-simulated hospitalizations attributable to COVID-19 in Years 1-4 at maximum likelihood parameter estimates, under four eligibility scenarios, assuming that vaccines reduce transmission potential of infectious individuals but not infection risk of susceptible individuals. Error bars represent the weekly 2.5 and 97.5 percentiles from 94 trajectories.

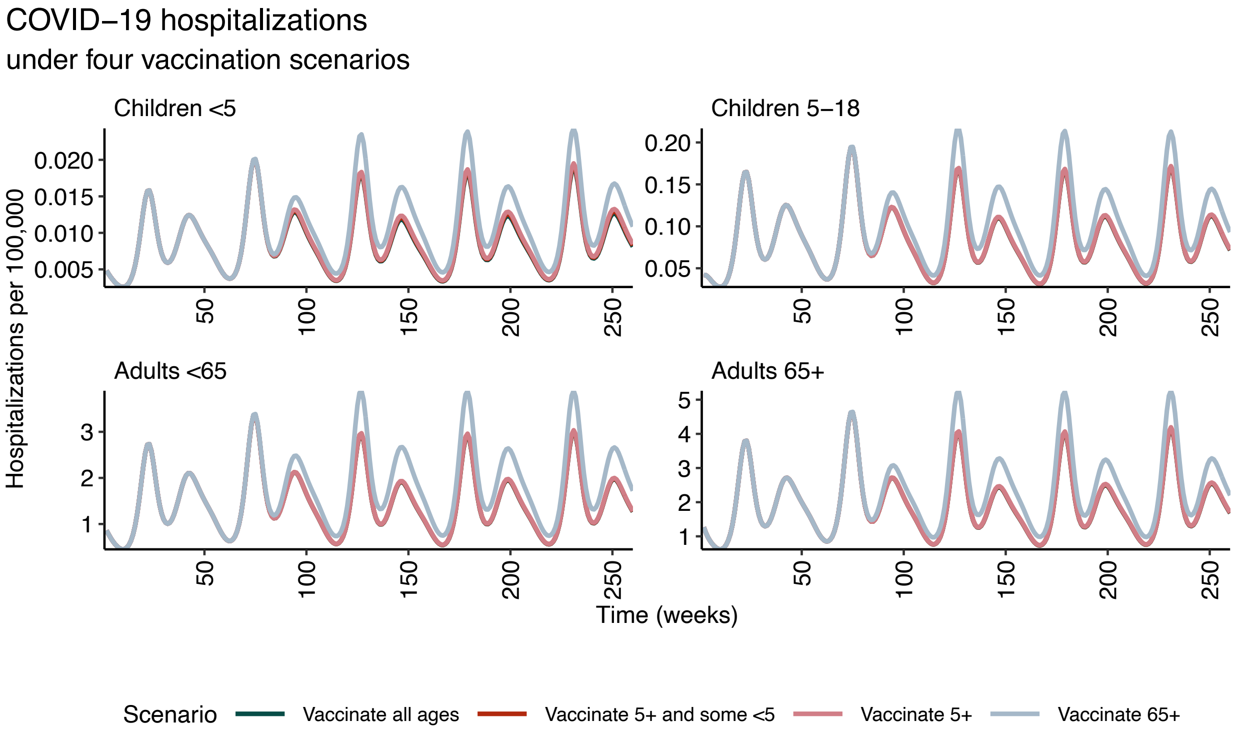


Figure S4: Age-stratified, model-simulated hospitalizations attributable to COVID-19 over Years 0-4 at maximum likelihood parameter estimates, under four eligibility scenarios, assuming that vaccines reduce transmission potential of infectious individuals but not infection risk of susceptible individuals.

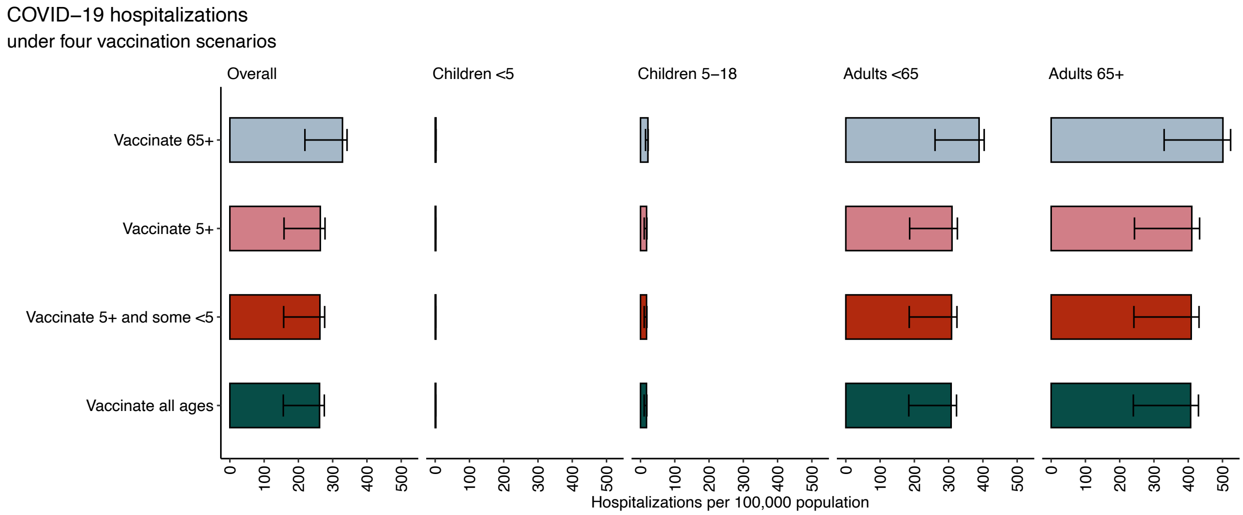


Figure S5: Alternate version of Figure S3 with uniform scale: model-simulated hospitalizations attributable to COVID-19 in Years 1-4 at maximum likelihood parameter estimates under four eligibility scenarios, assuming that vaccines reduce transmission potential of infectious individuals only. Error bars represent the weekly 2.5 and 97.5 percentiles from 94 trajectories.

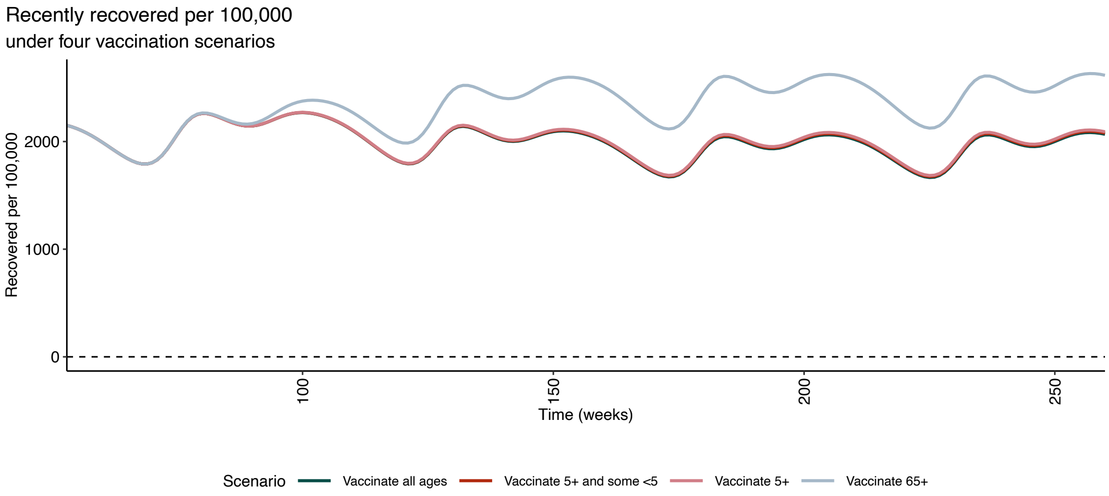


Figure S6: Recently recovered per $100,000$ in Years 1-4 , at maximum likelihood parameter estimates, assuming that vaccination reduces the transmission potential of infectious individuals. Three alternative eligibility scenarios are compared to the reference scenario with all-ages vaccination coverage. Error bars represent 2.5 and 97.5 percentiles of 94 paired trajectories, with parameters sampled from their 95% confidence intervals.

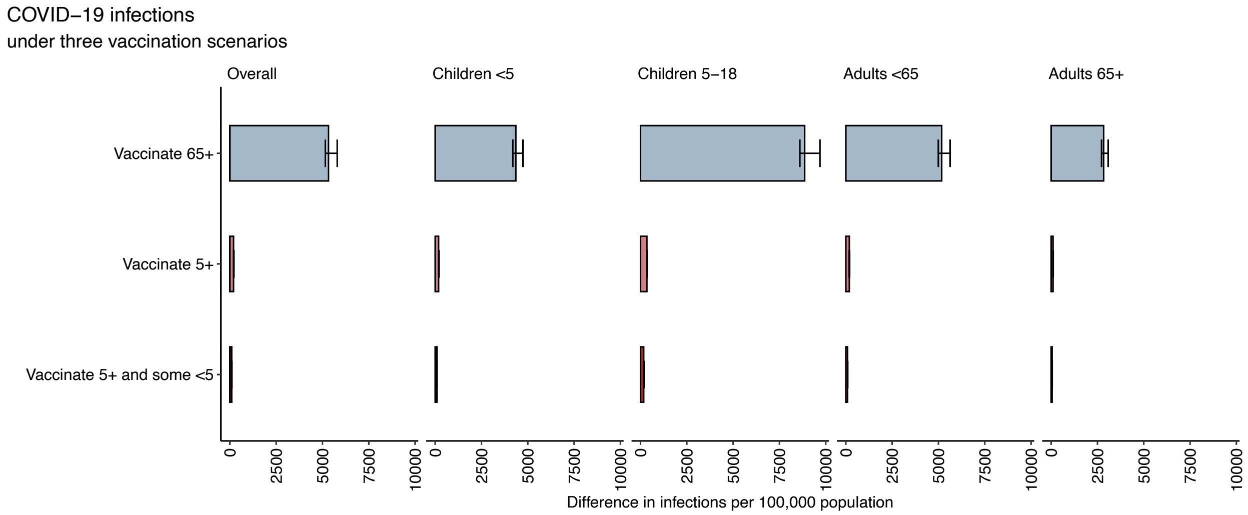


Figure S7: Difference in model-simulated infections attributable to COVID-19 in Years 1-4 , at maximum likelihood parameter estimates, assuming that vaccination reduces the transmission potential of infectious individuals. Three alternative eligibility scenarios are compared to the reference scenario with all-ages vaccination coverage. Error bars represent 2.5 and 97.5 percentiles of 94 paired trajectories, with parameters sampled from their 95% confidence intervals.

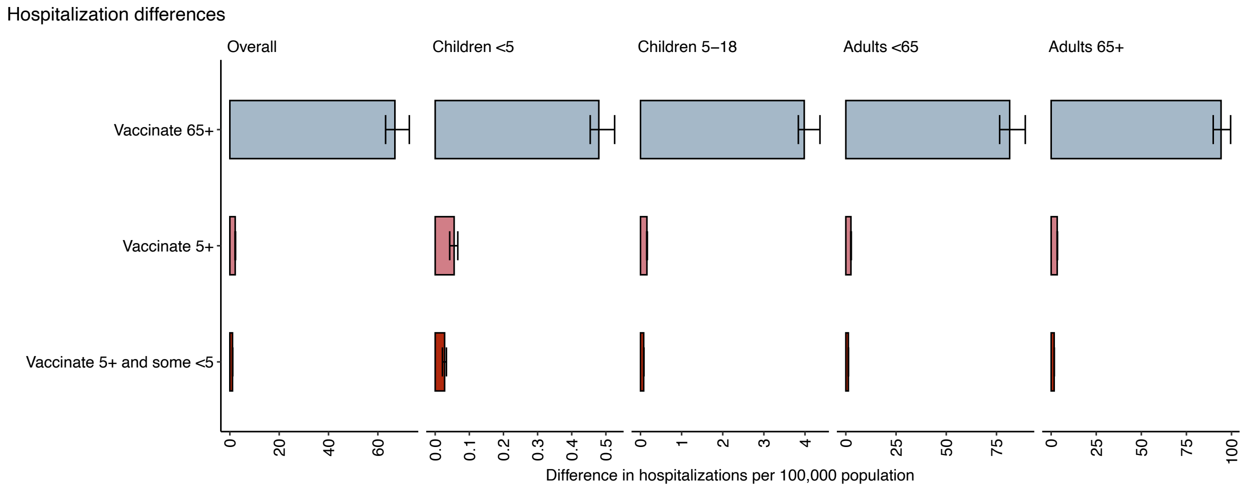


Figure S8: Difference in model-simulated hospitalizations attributable to COVID-19 over four years, at maximum likelihood parameter estimates, assuming that vaccination reduces the transmission potential of infectious individuals. Three alternative eligibility scenarios are compared to the reference scenario with all-ages vaccination coverage. Error bars represent 2.5 and 97.5 percentiles of 94 paired trajectories, with parameters sampled from their 95% confidence intervals.

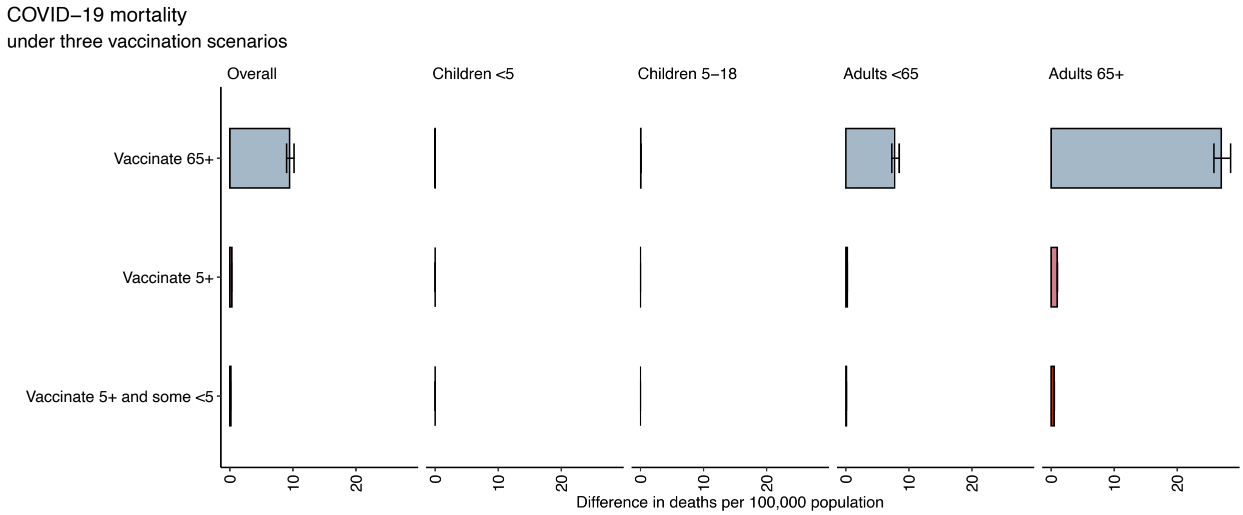


Figure S9: Difference in model-simulated deaths attributable to COVID-19 in Years 1-4, at maximum likelihood parameter estimates, assuming that vaccination reduces the transmission potential of infectious individuals. Three alternative eligibility scenarios are compared to the reference scenario with all-ages vaccination coverage. Error bars represent 2.5 and 97.5 percentiles of 94 paired trajectories, with parameters sampled from their 95% confidence intervals.

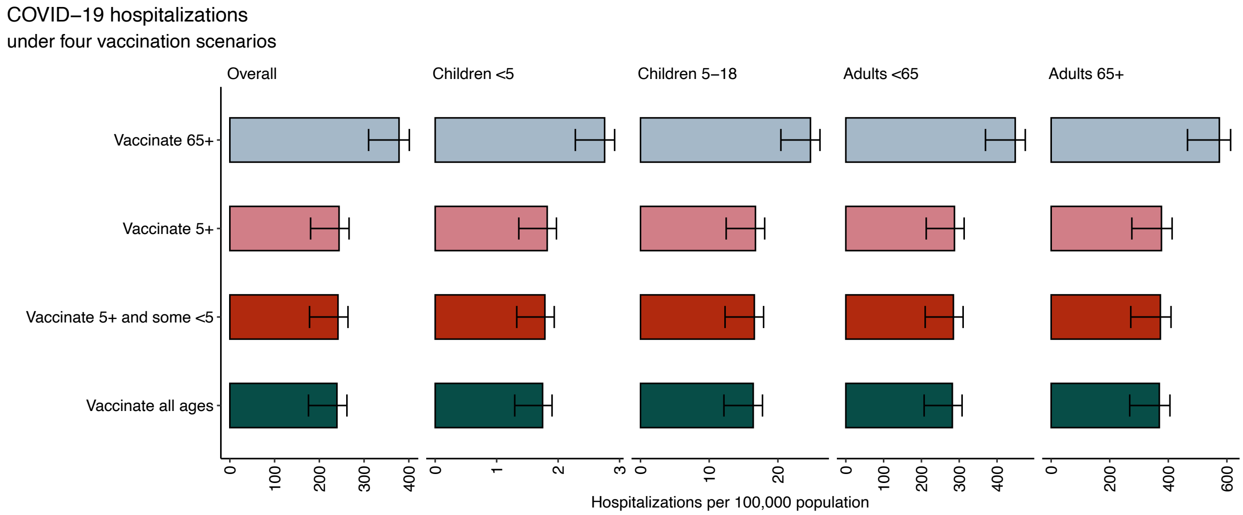


Figure S10: Model-simulated hospitalizations attributable to COVID-19 in Years 1-4 at maximum likelihood parameter estimates under four eligibility scenarios, assuming that vaccines reduce both infection risk for susceptible individuals and transmission potential for infectious individuals. Error bars represent the weekly 2.5 and 97.5 percentiles from 101 trajectories.

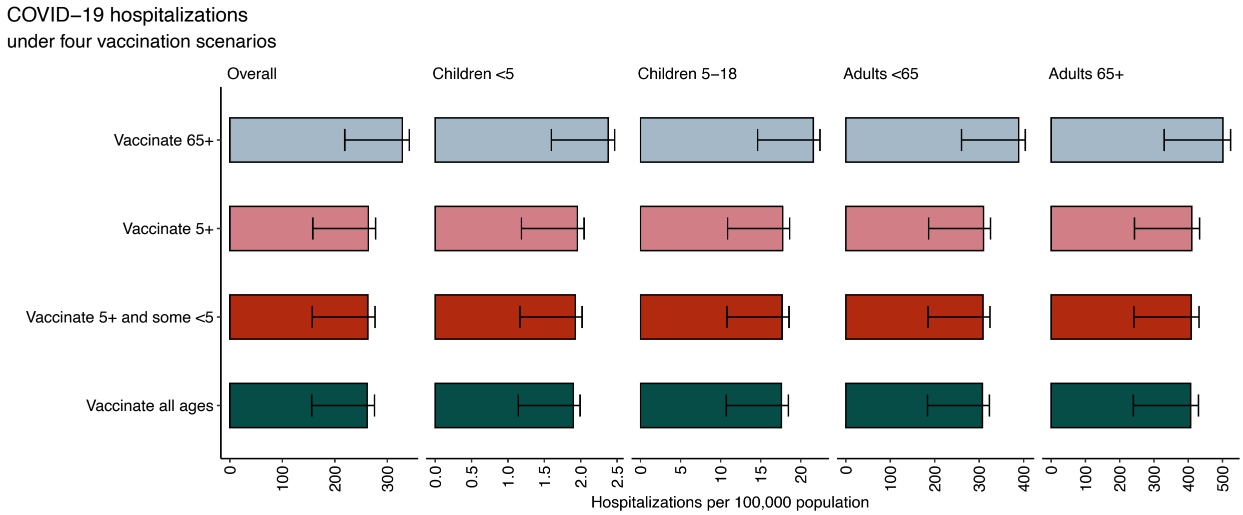


Figure S11: Model-simulated hospitalizations attributable to COVID-19 in Years 1-4 at maximum likelihood parameter estimates under four eligibility scenarios, assuming that vaccines reduce infection risk for susceptible individuals but not transmission potential for infectious individuals. Error bars represent the weekly 2.5 and 97.5 percentiles from 101 trajectories.

### Supplementary Tables

| Variable | – | Unweighted | Weighted |
|---|---|---|---|
| Wave | 1 | 3653 | 3646.51 |
| | 2 | 5310 | 5316.49 |
| Gender | Female | 5262 | 5132.79 |
| | Trans female/trans woman | 11 | 8.66 |
| | Male | 3619 | 3750.64 |
| | Trans male/trans man | 19 | 21.11 |
| | Non-binary / Gender nonconforming | 42 | 42.48 |
| | Other | 9 | 6.69 |
| | No response | 1 | 0.63 |
| Age | 19-29 | 1223 | 1553.59 |
| | 30-39 | 1583 | 1553.59 |
| | 40-49 | 1756 | 1553.59 |
| | 50-59 | 1558 | 1434.08 |
| | 60-69 | 1587 | 1434.08 |
| | 70-79 | 1080 | 956.05 |
| | 80+ | 176 | 478.03 |
| Income | Under $50,000 | 3224 | 2826.17 |
| | $50-$100,000 | 2702 | 2521.12 |
| | $100-$150,000 | 1499 | 1623.93 |
| | $150,000 or more | 1538 | 1991.78 |
| Education | Less than a high school education | 173 | 850.63 |
| | High school or equivalent | 1651 | 2265.37 |
| | Some college | 2766 | 2417.59 |
| | Bachelor's degree | 2490 | 2041.52 |
| | Postgraduate education | 1883 | 1387.88 |
| Race and ethnicity | White | 5572 | 5172.68 |
| | Hispanic/Latino | 1062 | 1694.5 |
| | Black or African American | 1480 | 1159.39 |
| | Asian | 447 | 535.1 |
| | Multiracial | 252 | 356.74 |
| | Other | 150 | 44.59 |
| Political affiliation | Republican | 2477 | 2590.89 |
| | Independent | 2005 | 2024.78 |
| | Democrat | 3620 | 3379.61 |
| | Other | 834 | 925.8 |
| | No response | 27 | 41.92 |

Table S1: Weighted and unweighted survey sample characteristics.

| Vaccine Mechanism | Compartment | 0-4 years | 5-18 years | 19-64 years | 65+ years |
|---|---|---|---|---|---|
| Infection blocking | S | 611132.25 | 1909344.17 | 6914675.64 | 1795696.94 |
| | SV | 9013 | 84848.39 | 490588.18 | 344412.14 |
| | I | 611.68 | 1910.46 | 6825.15 | 1728.26 |
| | IV | 9.02 | 84.9 | 485.4 | 333.74 |
| | H | 0.06 | 0.79 | 96.45 | 69.24 |
| | HV | 0 | 0.03 | 5.68 | 11.02 |
| | R | 32197.05 | 100592.39 | 364294.59 | 94604.97 |
| | RV | 474.84 | 4470.17 | 25846.28 | 18145.1 |
| Transmission blocking | S | 611132.25 | 1909344.17 | 6914675.64 | 1795696.94 |
| | SV | 9013 | 84848.39 | 490588.18 | 344412.14 |
| | I | 611.68 | 1910.46 | 6825.15 | 1728.26 |
| | IV | 9.02 | 84.91 | 487.32 | 337.45 |
| | H | 0.06 | 0.79 | 96.45 | 69.24 |
| | HV | 0 | 0.02 | 3.76 | 7.3 |
| | R | 32197.05 | 100592.39 | 364294.59 | 94604.97 |
| | RV | 474.84 | 4470.17 | 25846.28 | 18145.1 |
| Transmission and infection blocking | S | 611132.25 | 1909344.17 | 6914675.64 | 1795696.94 |
| | SV | 9013 | 84848.39 | 490588.18 | 344412.14 |
| | I | 611.68 | 1910.46 | 6825.15 | 1728.26 |
| | IV | 9.02 | 84.9 | 485.4 | 333.74 |
| | H | 0.06 | 0.79 | 96.45 | 69.24 |
| | HV | 0 | 0.03 | 5.68 | 11.02 |
| | R | 32197.05 | 100592.39 | 364294.59 | 94604.97 |
| | RV | 474.84 | 4470.17 | 25846.28 | 18145.1 |

Table S2: Initial conditions at the start of simulations. Omitted compartments are 0 at the start of simulations.

| Parameter | Meaning | Estimate | Source |
|---|---|---|---|
| Latent period | $1/\phi$ | 3 days | [? ] |
| Infection hospitalization rate (0-4) | $\eta_{0-4}$ | 0.0001 | [20] |
| Infection hospitalization rate (5-18) | $\eta_{5-18}$ | 0.0004 | [20] |
| Infection hospitalization rate (19-64) | $\eta_{19-64}$ | 0.0139 | [20] |
| Infection hospitalization rate (65+) | $\eta_{65+}$ | 0.0385 | [20] |
| Infectious period | $1/\gamma$ | 5 days | – |
| Hospitalization fatality rate (0-4) | $\sigma_{0-4}$ | 0.0040 | [8, 20] |
| Hospitalization fatality rate (5-18) | $\sigma_{5-18}$ | 0.0110 | [8, 20] |
| Hospitalization fatality rate (19-64) | $\sigma_{\text{19-64}}$ | 0.0953 | [8, 20] |
| Hospitalization fatality rate (65+) | $\sigma_{65+}$ | 0.2875 | [8, 20] |
| Duration of infection-induced immunity | $1/\omega$ | 8 months | – |
| Duration of vaccine-induced immunity | $1/\omega_v$ | 12 months | – |
| Vaccine efficacy against transmission | $\tau$ | 0%, 22% | [27] |
| Vaccine efficacy against infection | $VE_i$ | 0%, 34% | [? ] |
| Vaccine efficacy against severe disease given infection | $VE_s$ | 45%, 17% | [25] |
| Aging (0-4) | $\alpha_{0-4}$ | 5 years | |
| Aging (5-18) | $\alpha_{5-18}$ | 13 years | |
| Aging (19-64) | $\alpha_{19-64}$ | 47 years | |
| Aging (65+) | $\alpha_{65+}$ | 15 years | |

Table S3: Parameters used in model simulations.

| Age | Surveyed vaccination coverage |
|---|---|
| 0-4 | 5.81% |
| 5-18 | 17.02% |
| 19-64 | 26.50% |
| 65+ | 64.37% |

Table S4: Reported vaccination coverage from the Illinois survey (weighted).

| Age | Initialized vaccination coverage |
|---|---|
| 0-4 | 1.45% |
| 5-18 | 4.25% |
| 19-64 | 6.62% |
| 65+ | 16.09% |

Table S5: Initial vaccination coverage in model simulations, scaled from the Illinois survey.

| Age | $W_h$ | $V_m$ | $k$ |
|---|---|---|---|
| 0-4 | 18.33 | 0.07 | 4.87 |
| 5-18 | 17.66 | 0.05 | 5.02 |
| 19-64 16.24 | 0.09 | 4.62 | |
| 65+ | 14.69 | 0.35 | 5.19 |

Table S6: Type-III functional response parameters, calibrated from [23].

| Age | $V_m$ | $V_m$ (5+) | $V_m$ (65+) | $V_m$ (5+ and some $< 5$) |
|---|---|---|---|---|
| 0-4 | 0.07 | 0 | 0 | 0.03 |
| 5-18 | 0.05 | 0.05 | 0 | 0.05 |
| 19-64 | 0.09 | 0.09 | 0 | 0.09 |
| 65+ | 0.35 | 0.35 | 0.35 | 0.35 |

Table S7: Type-III functional response parameters, calibrated from [23].

| Parameter | Estimate | 2.5 | 97.5 |
|---|---|---|---|
| $\mu_1$ | 0.10 | 0.08 | 0.11 |
| $\mu_2$ | 0.11 | 0.078 | 0.13 |
| $\mu_3$ | 0.08 | 0.068 | 0.11 |
| $\mu_4$ | 0.17 | 0.148 | 0.24 |
| $\mu_5$ | 0.05 | 0.038 | 0.07 |
| $\mu_6$ | 0.14 | 0.13 | 0.20 |

Table S8: Maximum likelihood estimates and 95% confidence intervals for seasonal transmission magnitude parameters, assuming that vaccines both reduce infection risk for susceptible individuals and reduce transmission potential of infectious individuals.

| Parameter | Estimate | 2.5 | 97.5 |
|---|---|---|---|
| $\mu_1$ | 0.10 | 0.08 | 0.11 |
| $\mu_2$ | 0.10 | 0.07 | 0.13 |
| $\mu_3$ | 0.08 | 0.06 | 0.11 |
| $\mu_4$ | 0.17 | 0.14 | 0.24 |
| $\mu_5$ | 0.05 | 0.03 | 0.07 |
| $\mu_6$ | 0.14 | 0.12 | 0.20 |

Table S9: Maximum likelihood estimates and 95% confidence intervals for seasonal transmission magnitude parameters, assuming that vaccines reduce infection risk for susceptible individuals.

| Parameter | Estimate | 2.5 | 97.5 |
|---|---|---|---|
| $\mu_1$ | 0.09 | 0.08 | 0.11 |
| $\mu_2$ | 0.10 | 0.07 | 0.13 |
| $\mu_3$ | 0.08 | 0.06 | 0.11 |
| $\mu_4$ | 0.16 | 0.14 | 0.24 |
| $\mu_5$ | 0.05 | 0.03 | 0.07 |
| $\mu_6$ | 0.14 | 0.12 | 0.20 |

Table S10: Maximum likelihood estimates and 95% confidence intervals for seasonal transmission magnitude parameters, assuming that vaccines reduce transmission potential of infectious individuals.
